# The Social and Economic Factors Underlying the Incidence of COVID-19 Cases and Deaths in US Counties

**DOI:** 10.1101/2020.05.04.20091041

**Authors:** Nivedita Mukherji

## Abstract

This paper uncovers the socioeconomic and health/lifestyle factors that can explain the differential impact of the coronavirus pandemic on different parts of the United States. Using a dynamic panel representation of an epidemiological model of disease spread, the paper develops a Vulnerability Index for US counties from daily reported number of cases over a 20-day period of rapid disease growth. County-level economic, demographic, and health factors are used to explain the differences in the values of this index and thereby the transmission and concentration of the disease across the country. These factors are also used to examine the number of reported deaths. The paper finds that counties with high median income have a high incidence of cases but reported lower deaths. Income inequality as measured by the Gini coefficient, is found to be associated with more deaths and more cases. The remarkable similarity in the distribution of cases across the country and the distribution of distance-weighted international passengers served by the top international airports is evidence of the spread of the virus by way of international travel. The distributions of age, race, and health risk factors such as obesity and diabetes are found to be particularly significant factors in explaining the differences in mortality across counties. Counties with better access to health care as measured by the number of primary care physicians per capita have lower deaths, and so do places with more health awareness as measured by flu vaccination prevalence. Environmental health conditions such as the amount of air pollution is found to be associated with counties with higher deaths from the virus. It is hoped that research such as these will help policymakers to develop risk factors for each region of the country to better contain the spread of infectious diseases in the future.

## 1 Introduction

The novel coronavirus brought the global economy to a screeching halt in 2020 as it swept through much of the globe. The United States (US) was taken by surprise as it took the lead in not only the total number of positive cases but in terms of the number of reported deaths as well. By April, the number of cases for a single state such as New York exceeded the total number reported for any country, including China.

This paper attempts to uncover the socioeconomic conditions that are dominant in the areas that were most significantly impacted during the initial period of growth of the virus in the US. The literature on the transmission of infectious diseases often finds that highest impact areas have low income, poor sanitary conditions, and poor health care conditions due to their focus on viruses that have significantly impacted developing countries (Campos et al. (2018) for Zika, Redding et al. (2019) for Ebola are recent examples). Moore et al. (2017) used Ebola to develop an Infectious Disease Vulnerability Index for countries in Africa. The literature on the socioeconomic determinants of the spread of infectious diseases in developed countries is not extensive - Adda (2016) is an exception. Using data from France, it offers an extensive analysis of the transmission of three viruses - influenza, gastroenteritis, and chickenpox. The paper asks the important questions whether the virus spread more rapidly during periods of economic growth and if their spread follows a “gradient determined by economic factors.” Using data from France, Adda (2016) finds that the viruses studied propagated faster during times of economic boom due to increased economic activity and contact between people. Qiu (2020) have conducted a similar analysis for Wuhan, China. Both papers find a positive relationship between the spread of the virus and economic activity. Avery et al. (2020) offers a list of resources both in terms of relevant research and data sources for researchers.

Unlike some of the papers cited above that concentrate on the impact of mitigation and/or containment strategies along with economic conditions such as GDP, employment, and weather-related factors such as temperatures, and pollution (Wu (2020)), this paper focuses on economic, demographic, and health conditions in explaining the number of cases and deaths in the US during its peak growth period. Figure 1 clearly indicates that the spread of the virus has been in regions of high economic activity on the two coasts. The virus arrived on the US shores through international travel. While the initial spread of the virus is expected to be triggered by international travel and economic activity, it is important to understand whether its continued spread and concentration are restricted to such places. This paper attempts to understand the underlying socioeconomic conditions of the geographic regions around the US that made them susceptible to becoming hotspots. This is related to the question about the factors that determine the gradient followed by the virus as it spreads through the country.

**Figure 1:**
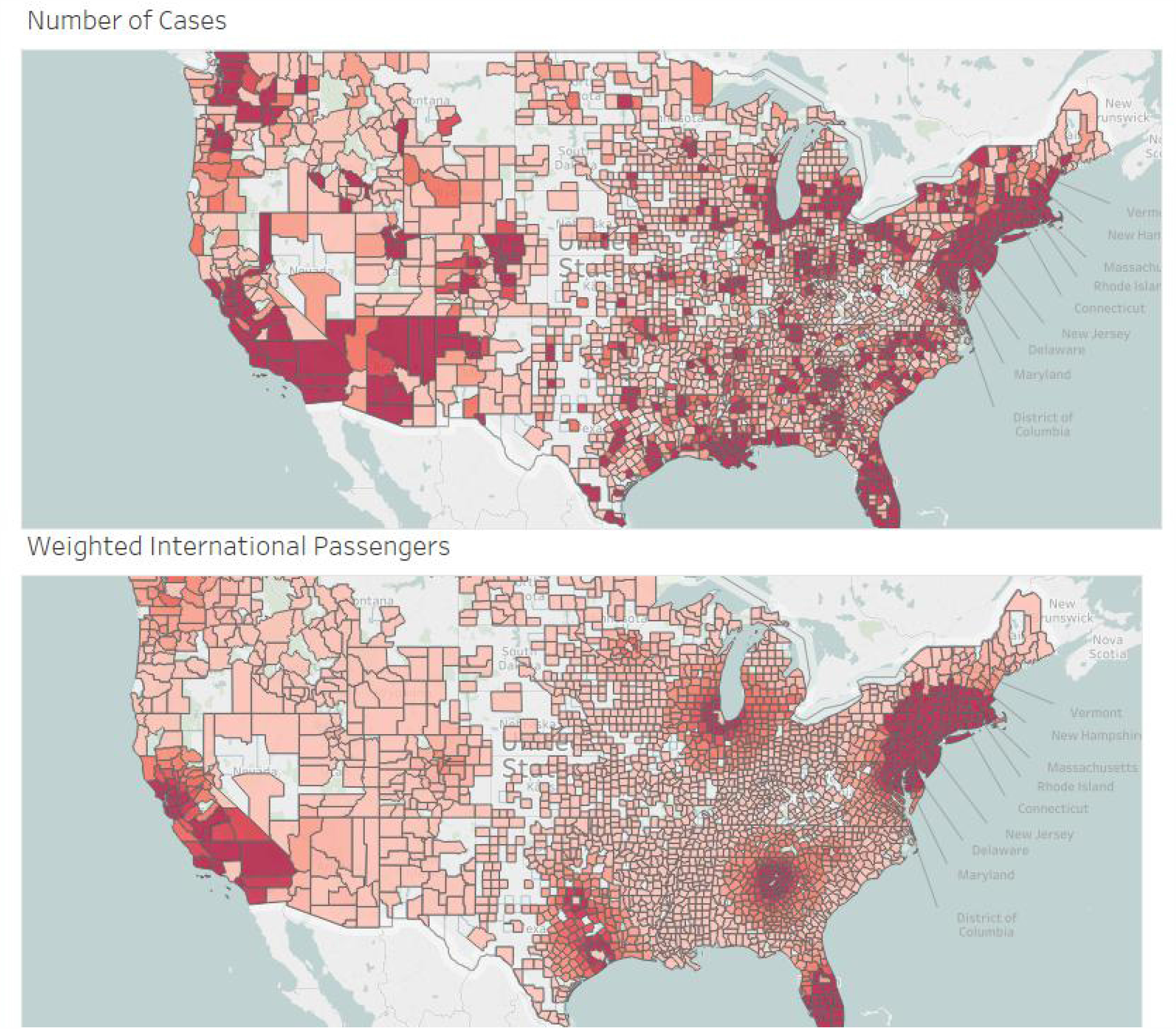
Distribution of Cases and International Passengers.

Introducing heterogeneity that captures region-specific uniqueness in an epidemiological model of disease spread, the paper develops a Vulnerability Index for the counties included in the study. The index captures the underlying factors that impact the vulnerability of a region to the virus. Economic, demographic, and health/lifestyle factors are used to explain the observed differences in the vulnerability index. These factors are also used to explain the differences in the number of deaths reported across the counties. Data used for the analysis is obtained from the end of March to April 20, 2020. The reason for the selection of this period is due to the fact that this phase witnessed the most rapid rise of the infection in the US. To prepare for future infectious diseases, it is important to be able to better predict if the population of a certain area is particularly vulnerable to such a disease. This paper shows that it is possible to develop a vulnerability index for disease spread based on the socioeconomic composition of the population and their health/lifestyle choices. Developing such a profile will help policymakers to determine the places that need to be protected first when the threat of such a disease appears on the horizon.

Recent experience suggests that infectious diseases are a major threat to both the health and economic well being of people around the world. In spite of the experience with H1N1, SARS, and Ebola, countries such as the US did not develop a coherent infrastructure or strategy to determine which parts of the country are at a particularly higher risk of disease transmission. This paper shows that it is possible to utilize the economic, demographic, and lifestyle profiles of regions to develop a risk factor for each geographical area so that when the next epidemic arises, public officials are better prepared to anticipate where the hotspots are likely to arise and take the necessary containment steps. The experience with the coronavirus shows how rapidly an infectious disease can bring an economy down. Without advance preparation, the next disease will be just as difficult to contain as this. The large differences within state boundaries show the importance of developing more local strategies that take into consideration a multitude of factors.

## 2 Methodology and Data

The coronavirus pandemic impacted all 50 states in the US. The experience of each state, county, and city has been anything but homogeneous. To understand this differential effect across counties in the US, we consider two sets of factors - epidemiological factors that explain the spread of infectious diseases and socioeconomic factors that enhance or mitigate the spread of the disease. Epidemiological models explain how an infectious disease evolves in a region based on population and the size of the pool of infected individuals. We will use epidemiological models such as the SIR model to determine the fundamental differences in cases based on population size and the number of infections. These factors alone cannot explain the entire heterogeneous outcomes across the country. We expect differences in types and amounts of economic activities, living conditions, demographic makeups, and lifestyle choices to determine the vulnerabilities of communities in the spread of a highly contagious virus such as the coronavirus.

We will conduct this analysis in two steps. In the first step, an epidemiological model of disease spread will be used to generate estimates of a vulnerability index for each county once population and infections are accounted for. In the second step, we will use county-level economic, demographic, and health data to explain differences in the vulnerability indexes across counties.

Epidemiological models of the SIR type such as in Blackwood et al. (2018) describe disease spread dynamics based on three main factors - the size of the population, the number of susceptible individuals, and the number of infected individuals. With a population of size *N*, if *I* denotes the number of infected individuals, *R* denotes the number of recovered individuals, the number of individuals susceptible to the disease is given by *S* = *N* − *I* − *R*. At each time *t*, the number of new infections will depend on the interactions of the susceptible (*S*) and infected (*I*) individuals. The infected individuals are non-infectious during the latent period and asymptomatic but infectious from the end of the latent period to the end of the incubation period and infectious with symptoms after the end of the incubation period. If *j* denotes the number of days it takes to become infectious, at time *t*, the interactions of susceptible people with people infected *t* − *j* days earlier will lead to new cases.

Using daily reports of coronavirus cases for counties across the US, we generate a panel dataset of US counties over a 20 day period from March 30 to April 18. This period captures the period of the rapid growth of cases in the US. The panel data approach in estimating the growth of the virus in different parts of the US allows us to introduce county-specific fixed effects in the estimation. The panel estimates the number of cases as a function of the potential pool of susceptible and infected individuals and time and county-specific fixed effects and is given by the following equation:

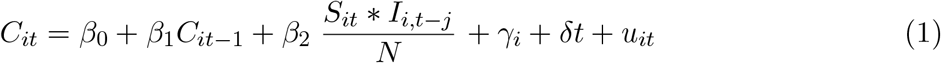

where, *C*^*it*^ denotes the number of reported cases in county *i* at time *t, γ*^*i*^ gives the fixed effect parameter for county *i, δ* is the parameter for the time variable, and *u*^*it*^ is the error for county *i* at time *t*.

Estimation of the above regression will generate the county fixed-effect value, *γ*, for each county. From these fixed effects, we will generate a vulnerability index for each county. This approach is similar to the one used by Mukherji and Silberman (2013) in studying patent citations between metro areas in the US.

In the second step of the analysis, we will use county-level economic and demographic factors to explain how they influence the vulnerability index for each county. The economic factors include income, unemployment rate, income inequality, and access to housing. The set of demographic factors include the size of the population and its density, the racial profile of the counties, the age distribution of the population, and the percentage of the population that was born outside the US. In addition to the county level economic and demographic data, spatial factors are considered as well. The contagious nature of the disease compels one to consider the spillover effects on neighboring counties. We introduce inverse-distance weighted values of the number of international passengers served by the top 46 international airports in the contiguous US. Since the virus is presumed to have originated in China and then spread to other parts of the world, including Europe, before taking hold in the US, international passenger data is introduced to examine if proximity to international airports is related to the concentration of confirmed cases. While international passengers often arrive at a particular airport and then use domestic airlines to travel to other parts of the country, the locations of the international airports are closely tied to areas with concentrations of activities that are globally oriented. Consequently, the international passengers served by these airports are expected to interact in the regions around these airports in large numbers. Using a 300-mile radius around each county where the airports are located, an inverse-distance matrix is used to assign the number of international passengers in the areas surrounding the airports. The bottom part of Figure 1 displays the weighted distribution of international passengers. While this data is unrelated to the number of confirmed cases, the spatial distribution of the passenger data is similar to the spatial distribution of confirmed cases.

The estimation of the impact of these regional factors in explaining differences in vulnerabilities to the disease will be based on Equation (2).

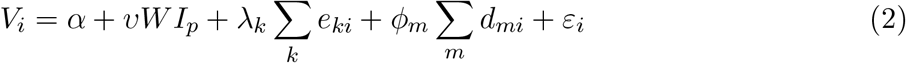

In the above equation, *V*^*i*^ represents the vulnerability index of county *i, e*^*ki*^ represents the set of *k* economic variables that makes a county susceptible to the spread of the disease due to the enhanced interactions between people and working in close proximity. Although the economic activity of a county changes with time, the general distribution of such activities across the country remains relatively stable within short periods of time; *d*^*m*^ represents the demographic factors. This equation includes the spatially weighted number of international passengers in a region by multiplying an inverse distance-weighted matrix W with the number of international passengers, *I*, served by an international airport in the neighborhood of county *i*. The data and estimation results are reported in the next section.

## 3 Estimation

### 3.1 Data

The data on COVID-19 cases and deaths is obtained from the COVID tracking data provided by the New York Times and Johns Hopkins University. Figure 1 displays the distribution of cases in the 2512 counties.

Data for various demographic and economic variables such as population distribution by ethnicity, population density are obtained from data compiled by the USDA’s Atlas of Rural and Small Town America and the Federal Communication Commission. The underlying data come from the Census Bureau and the American Medical Association. Some of the demographic data, such as the distribution of the population by race, are from the 2010 census. The total population, per capita personal income, unemployment data are from 2018. The percentage of the population with various health-related factors such as obesity and diabetes are available from the 2014-18 period. The Wisconsin County Health Rankings data are used for data on flu vaccination and air pollution. Data on international air passengers were obtained from the Bureau of Transportation Statistics. This source provides the number of international passengers served by the top 50 international airports in the US. Using airports in the contiguous US only, 46 of the 50 airport data were used. The total number of passengers on international flights was over 109 million in 2018. In order to account for local spillover effects of the virus in the form of increased susceptibility due to a higher prevalence of cases, an inverse distance weighted matrix was created with positive weights assigned up to a 300-mile radius around a county. This radius is just large enough to ensure that each county in the study had at least one other county in the study as a neighbor.

### 3.2 Estimation of Cases

The previous section explained that the foundation of the analysis of the socioeconomic factors that can contribute to the spread and concentration of the coronavirus in the various parts of the country lies in the epidemiological model of disease transmission. The first step is to generate county-level vulnerability measures from an estimation of equation (1). The daily coronavirus data is available for over 2500 counties. To manage the computational load of estimating a panel that large, we restrict our analysis to counties that reported an average of 30 cases per day from March 30 through April 19. This generates a panel of 771 counties covering all 50 states. Each of the counties reported at least one confirmed case during the period of analysis resulting in a balanced panel. Equation (1) includes a lagged value of infections in determining the proportion of the population that is susceptible at any time *t*. The incubation period for this virus is estimated to be anywhere between 2 to 14 days. People are infections a few days before they develop symptoms and after they develop symptoms. We assume a seven day lag for the results reported in the paper. Sensitivity analysis was conducted for different lag lengths.

Since cases in period *t* depend on the number of cases in period *t* − 1, the estimation of equation (1) requires the use of dynamic panel estimation methods. A model with small T (20) and large N (771) with a lagged dependent variable is expected to have the Nickell’s bias Stephen (1981). A difference GMM estimation is found to be the best option for the data. The Allerano-Bond estimation method Arellano and Bond (1991) that uses lagged values as instruments as implemented by Roodman (2006) was used. The results are reported in Table 1 and show that although autocorrelation of the first order exists, there is no second-order autocorrelation. The Sargan and Hansen tests of no overidentification of instruments are satisfied, and the F statistic shows that the model fits the data well. The table shows that the one period lagged number of cases has a significant impact on the number of cases reported on any day. The interaction of the infected and susceptible population is also significant and positive.

**Table 1:** Dynamic Panel Regression of Disease Spread

| Variable | Coefficient | St.Err. | t-value | p-value | [95% Conf | Interval] |
| --- | --- | --- | --- | --- | --- | --- |
| $\ln Cases_{t-1}$ | 0.883*** | 0.051 | 17.280 | 0.000 | 0.783 | 0.983 |
| $\frac{\ln SI_{t-7}}{N}$ | 0.039* | 0.022 | 1.790 | 0.075 | -0.004 | 0.083 |
| D1-D8 | Omitted |  |  |  |  |  |
| D9 | -0.019*** | 0.008 | -2.470 | 0.014 | -0.034 | -0.004 |
| D10 | -0.016 | 0.010 | -1.640 | 0.102 | -0.034 | 0.003 |
| D11 | -0.019 | 0.012 | -1.580 | 0.115 | -0.042 | 0.005 |
| D12 | -0.030** | 0.014 | -2.180 | 0.030 | -0.057 | -0.003 |
| D13 | -0.037** | 0.016 | -2.330 | 0.020 | -0.068 | -0.006 |
| D14 | -0.030* | 0.016 | -1.820 | 0.070 | -0.062 | 0.002 |
| D15 | -0.033* | 0.019 | -1.740 | 0.082 | -0.070 | 0.004 |
| D16 | -0.037* | 0.020 | -1.800 | 0.073 | -0.077 | 0.003 |
| D17 | -0.026 | 0.022 | -1.220 | 0.224 | -0.069 | 0.016 |
| D18 | -0.024 | 0.023 | -1.030 | 0.304 | -0.070 | 0.022 |
| D19 | -0.029 | 0.025 | -1.150 | 0.250 | -0.078 | 0.020 |
| D20 | -0.034 | 0.026 | -1.280 | 0.201 | -0.085 | 0.018 |
| *t* statistics in parentheses |  |  |  |  |  |  |
| * $p < 0.1$, ** $p < 0.05$, *** $p < 0.01$ | | | | | | |
| Arellano-Bond test for AR(1) in first differences: $z = -8.10$ $\Pr > z = 0.000$ | | | | | | |
| Arellano-Bond test for AR(2) in first differences: $z = 1.56$ $\Pr > z = 0.119$ | | | | | | |
| Sargan test of overid. restrictions: $\chi^2(18) = 15.15$ | | | | | Prob > $\chi^2 = 0.652$ | |
| Hansen test of overid. restrictions: $\chi^2(18) = 28.18$ | | | | | Prob > $\chi^2 = 0.059$ | |
| Group variable: Fips |  |  |  |  | Number of Obs = 10794 |  |
| Time variable : Day |  |  |  |  | Number of Groups = 771 |  |
| Number of instruments = 44 |  |  |  |  | Obs per Group: Min = 14 |  |
| F(22, 771) = 6006.10 |  |  |  |  | Avg = 14.00 |  |
| Prob > F = 0.000 |  |  |  |  | Max = 14 |  |

One of the key objectives of this regression is to obtain a set of estimates for the county level fixed effects. The method of dynamic panel estimation, such as GMM that utilizes first differencing, removes the impact of time-invariant variables such as the time-invariant fixed effects. These are, however, recoverable from the residuals. It is to be noted that for a dynamic panel model of the form, *y*_*it*_ = *ρy*_*it*−1_ + *a*_*i*_ + *e*_*it*_, the residual is given by 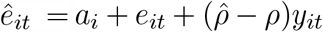. The average *ē*_*i*_ can be used as an estimator of the fixed effects to analyze how the underlying conditions in the various counties impact the fixed effects as long as those factors are uncorrelated with the *e*_*it*_. That condition is satisfied with the average *e*_*it*_ equaling −7.00e-09 for the results of the regression of equation 1.

### 3.3 Estimation of the Vulnerability Index

The estimates of the fixed effects derived from the dynamic panel regression of cases are converted to an index by transforming the mean value to 100 and is termed the Vulnerability Index. High values of the index indicate that the counties are more susceptible to the growth of the disease. The value of the index ranges from a low of 63 for Lincoln, Arkansas to a high of 229 for New York City (cases and deaths data is combined for the boroughs of New York City). Table 2 offers a list of the 20 lowest and highest values of the index. The results show that the higher values were in the so-called “hot spots”. The table lists the region codes and Urban Influence Codes (UIC) used by the USDA to distinguish between rural and urban areas. Codes 1 and 2 are for metro areas, 11 and 12 are for non-core areas that are not adjacent to any metro area. The table shows the concentration of the high index areas in the Northeast and in large metro areas. The bottom values are found in counties mainly outside the Northeast. There is a large difference in the population densities of the places with high values of the index than the ones with the smallest values. The table shows that there are differences in both location and type of county that distinguish areas with high values of infections from places with smaller outbreaks. We attempt to introduce additional factors that can shed light on why some places experienced significantly higher infection rates than others after controlling for the pool of susceptible individuals.

**Table 2:**
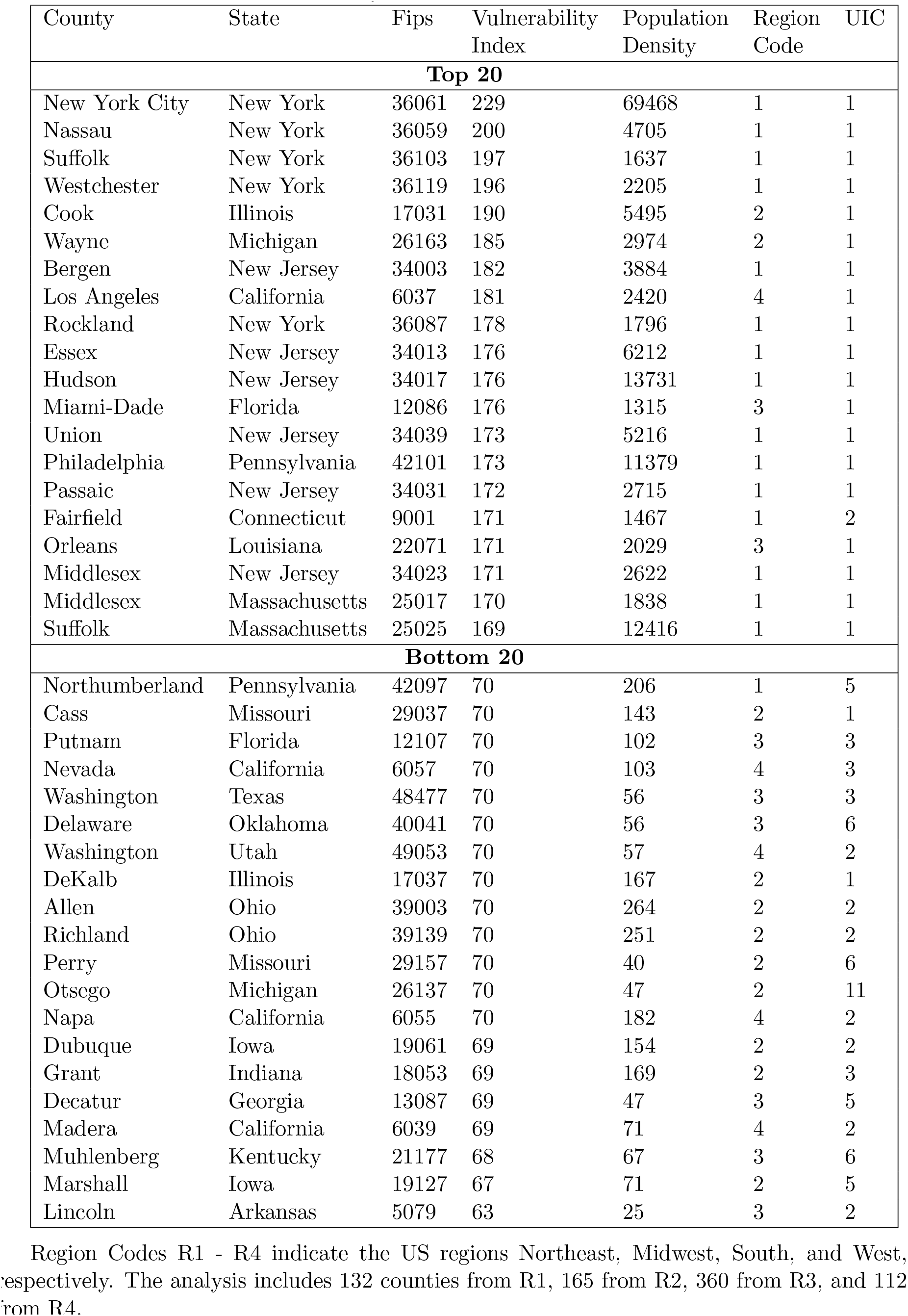
Vulnerability Index of Top and Bottom 20 Counties

| County | State | Fips | Vulnerability<br>Index | Population<br>Density | Region<br>Code | UIC |
| --- | --- | --- | --- | --- | --- | --- |
| **Top 20** |  |  |  |  |  |  |
| New York City | New York | 36061 | 229 | 69468 | 1 | 1 |
| Nassau | New York | 36059 | 200 | 4705 | 1 | 1 |
| Suffolk | New York | 36103 | 197 | 1637 | 1 | 1 |
| Westchester | New York | 36119 | 196 | 2205 | 1 | 1 |
| Cook | Illinois | 17031 | 190 | 5495 | 2 | 1 |
| Wayne | Michigan | 26163 | 185 | 2974 | 2 | 1 |
| Bergen | New Jersey | 34003 | 182 | 3884 | 1 | 1 |
| Los Angeles | California | 6037 | 181 | 2420 | 4 | 1 |
| Rockland | New York | 36087 | 178 | 1796 | 1 | 1 |
| Essex | New Jersey | 34013 | 176 | 6212 | 1 | 1 |
| Hudson | New Jersey | 34017 | 176 | 13731 | 1 | 1 |
| Miami-Dade | Florida | 12086 | 176 | 1315 | 3 | 1 |
| Union | New Jersey | 34039 | 173 | 5216 | 1 | 1 |
| Philadelphia | Pennsylvania | 42101 | 173 | 11379 | 1 | 1 |
| Passaic | New Jersey | 34031 | 172 | 2715 | 1 | 1 |
| Fairfield | Connecticut | 9001 | 171 | 1467 | 1 | 2 |
| Orleans | Louisiana | 22071 | 171 | 2029 | 3 | 1 |
| Middlesex | New Jersey | 34023 | 171 | 2622 | 1 | 1 |
| Middlesex | Massachusetts | 25017 | 170 | 1838 | 1 | 1 |
| Suffolk | Massachusetts | 25025 | 169 | 12416 | 1 | 1 |
| **Bottom 20** |  |  |  |  |  |  |
| Northumberland | Pennsylvania | 42097 | 70 | 206 | 1 | 5 |
| Cass | Missouri | 29037 | 70 | 143 | 2 | 1 |
| Putnam | Florida | 12107 | 70 | 102 | 3 | 3 |
| Nevada | California | 6057 | 70 | 103 | 4 | 3 |
| Washington | Texas | 48477 | 70 | 56 | 3 | 3 |
| Delaware | Oklahoma | 40041 | 70 | 56 | 3 | 6 |
| Washington | Utah | 49053 | 70 | 57 | 4 | 2 |
| DeKalb | Illinois | 17037 | 70 | 167 | 2 | 1 |
| Allen | Ohio | 39003 | 70 | 264 | 2 | 2 |
| Richland | Ohio | 39139 | 70 | 251 | 2 | 2 |
| Perry | Missouri | 29157 | 70 | 40 | 2 | 6 |
| Otsego | Michigan | 26137 | 70 | 47 | 2 | 11 |
| Napa | California | 6055 | 70 | 182 | 4 | 2 |
| Dubuque | Iowa | 19061 | 69 | 154 | 2 | 2 |
| Grant | Indiana | 18053 | 69 | 169 | 2 | 3 |
| Decatur | Georgia | 13087 | 69 | 47 | 3 | 5 |
| Madera | California | 6039 | 69 | 71 | 4 | 2 |
| Muhlenberg | Kentucky | 21177 | 68 | 67 | 3 | 6 |
| Marshall | Iowa | 19127 | 67 | 71 | 2 | 5 |
| Lincoln | Arkansas | 5079 | 63 | 25 | 3 | 2 |

Region Codes R1 - R4 indicate the US regions Northeast, Midwest, South, and West, respectively. The analysis includes 132 counties from R1, 165 from R2, 360 from R3, and 112 from R4.

Estimation of equation (2) sheds light on the factors that contribute most strongly to the differences in the values of the vulnerability index^1^ noted in Table 2. The estimation results are reported in Table 3. The results are reported for models that include population and population density separately and together. These two variables play an important role in disease transmission and have a correlation of 0.76. It is important to understand their individual and collective impact on the vulnerability index values of the counties. The other critical variables are classified into three broad groups - economic, demographic, and health/lifestyle. Since many of the conditions in a county are influenced by policies and conditions at the state level, we include a series of state dummy variables to control for state level influences.

**Table 3:** Regression Explaining Vulnerability Index

| Variable | Model 1 | Model 2 | Model 3 |
| --- | --- | --- | --- |
| **Economic Variables** |  |  |  |
| International Air Passengers | 0.00000785***<br>(2.74) | 0.00000577<br>(0.89) | 0.00000813***<br>(2.67) |
| Weighted International Air Passengers | 0.000207**<br>(2.23) | 0.0000382<br>(0.41) | 0.000212**<br>(2.24) |
| ln Median Income | 0.253***<br>(6.71) | 0.275***<br>(6.78) | 0.254***<br>(6.63) |
| Gini | 0.503**<br>(2.40) | 0.306<br>(1.24) | 0.508**<br>(2.40) |
| Severe Housing Problems | 0.00372*<br>(1.66) | 0.00441*<br>(1.76) | 0.00371*<br>(1.65) |
| **Demographic Variables** |  |  |  |
| ln Population | 0.134***<br>(19.98) |  | 0.136***<br>(16.19) |
| ln Population Density |  | 0.0867***<br>(11.99) | -0.00265<br>(-0.35) |
| % Population Over 65 | 0.00559***<br>(3.51) | 0.00316*<br>(1.90) | 0.00559***<br>(3.50) |
| % White | -0.00391***<br>(-6.43) | -0.00432***<br>(-6.91) | -0.00393***<br>(-6.32) |
| % Foreign Born | -0.000231<br>(-0.18) | 0.00301*<br>(1.91) | -0.000255<br>(-0.19) |
| **Community Health/Lifestyle Variables** |  |  |  |
| Preventable Hospital Stays Per 100000 | 0.000799**<br>(2.00) | 0.00125***<br>(2.81) | 0.000787**<br>(1.97) |
| ln Primary Care Physicians Per Capita | -0.00403<br>(-0.29) | 0.0162<br>(1.02) | -0.00383<br>(-0.28) |
| % Flu Vaccination | -0.000821<br>(-0.67) | 0.000882<br>(0.64) | -0.000797<br>(-0.65) |
| State Fixed Effects | Significant | Significant | Significant |
| Constant | -0.00385<br>(-0.01) | 1.256**<br>(2.46) | -0.00949<br>(-0.02) |
| Observations | 768 | 768 | 768 |
| $R^2$ | 0.772 | 0.699 | 0.772 |
| Adjusted $R^2$ | 0.752 | 0.673 | 0.752 |

$t$ statistics in parentheses
\* $p < 0.1$ , \*\* $p < 0.05$ , \*\*\* $p < 0.01$

The results show that in the economic group, median household income has a positive and significant effect showing that places of high income are more vulnerable. This is a reflection of the high human interactions that result from high economic activities and is consistent with the findings of Adda (2016). The Gini coefficient measuring the degree of income inequality and severe housing problems are both positive and significant, suggesting that places where large portions of the population experience poor economic, living, and housing conditions transmitted the virus more rapidly. Taken together, these results show that counties with high levels of economic activity but unequal distribution of income and living conditions are more vulnerable to the transmission of the disease.

Moreover, areas of high economic activities are also where the main international airports are located. The number of international passengers served by the airports is positive and significant in most models. This corroborates Figure 1 that showed that the locations of the international airports and the number of international passengers they serve overlap with the regions of high infection rates. The international passengers not only impact the counties in which the airports are located, their effects spill over to the neighboring regions as well. The results show that the distance-weighted number of international passengers served by these airports is positive and significant^2^.

The results related to the demographic variables are consistent with the findings from the health professionals. People living in counties with higher population and population density are more vulnerable since they lead to more interactions and hence transmission. Counties with a higher concentration of the elderly are more vulnerable, and so are places with a higher concentration of people of color. Places with a higher concentration of immigrants did not show any significant difference in vulnerability once population is included.

In addition to the economic and demographic factors that can contribute to a region’s vulnerability to the disease, we examine if some basic health-related factors play any role. We include three health-related variables - access to health care, individual choices to protect against bad health outcomes, and a measure of the population’s success in leading a healthy lifestyle. These are included to examine if the health behavior of the populations in the counties has any direct impact on their vulnerability to an infectious disease such as the coronavirus. Toward this goal, we include the number of primary care physicians per capita is included as an indicator of access to health care; the percentage of the population that receives the flu vaccine as a measure of the population’s voluntary health protection decisions; and the number of preventable hospital stays as an indicator of adverse health outcomes that result from poor health care decisions made by the people of a county. The most significant variable is found to be the preventable hospital stays variable showing that places where people’s adverse health care actions are harmful enough to lead to preventable hospital visits, are also places where people are more vulnerable to catching the coronavirus. Vulnerability is not found to be strongly related to having access to health care or taking preventive steps such as getting vaccinated.

The results reported in Table 3 show that collectively the economic, demographic, and to a lesser extent, the health care makeup of the counties can explain a large portion of the variance in county-level vulnerability index values. We consider next if these factors can also explain the differences in the number of deaths caused by Covid-19.

### 3.4 Estimation of Deaths

Table 4 displays the results of an estimation of the number of deaths by county. To avoid issues related to the dependent variable being counts of deaths and account for differences in population sizes, the data on deaths and cases are converted to values per 100,000 people. Since the rest of the independent variables are all expressed as percentages or values that do not require population adjustments, the transformation of the deaths and cases to values per 100,000 eliminates the need to include a control for population size. The data for deaths includes values for each of the 20 days of the study. We use a pooled regression methodology with indicator variables for the days for which the data are analyzed and US states to which the counties belong to control for time and same-state impacts.

**Table 4:**
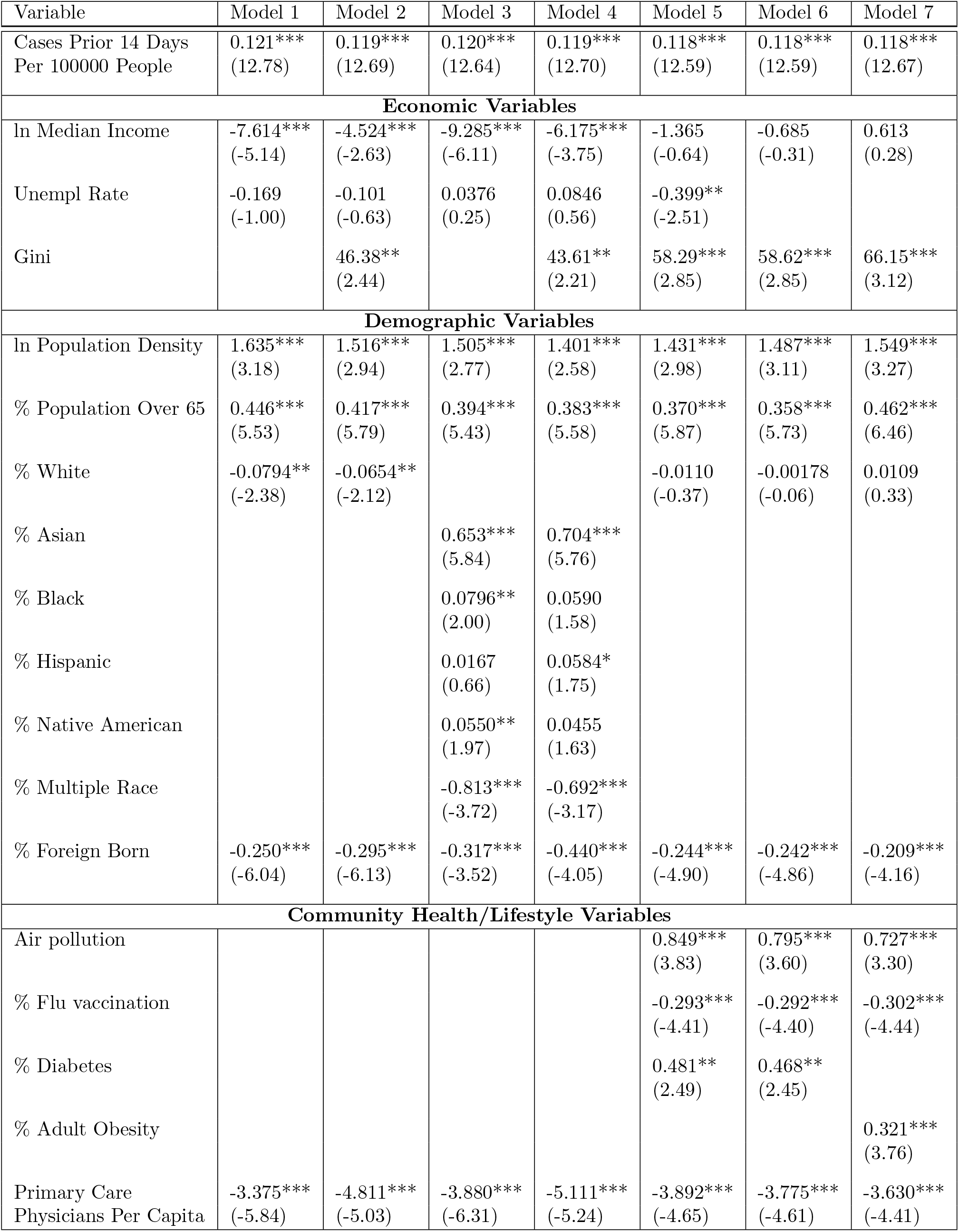
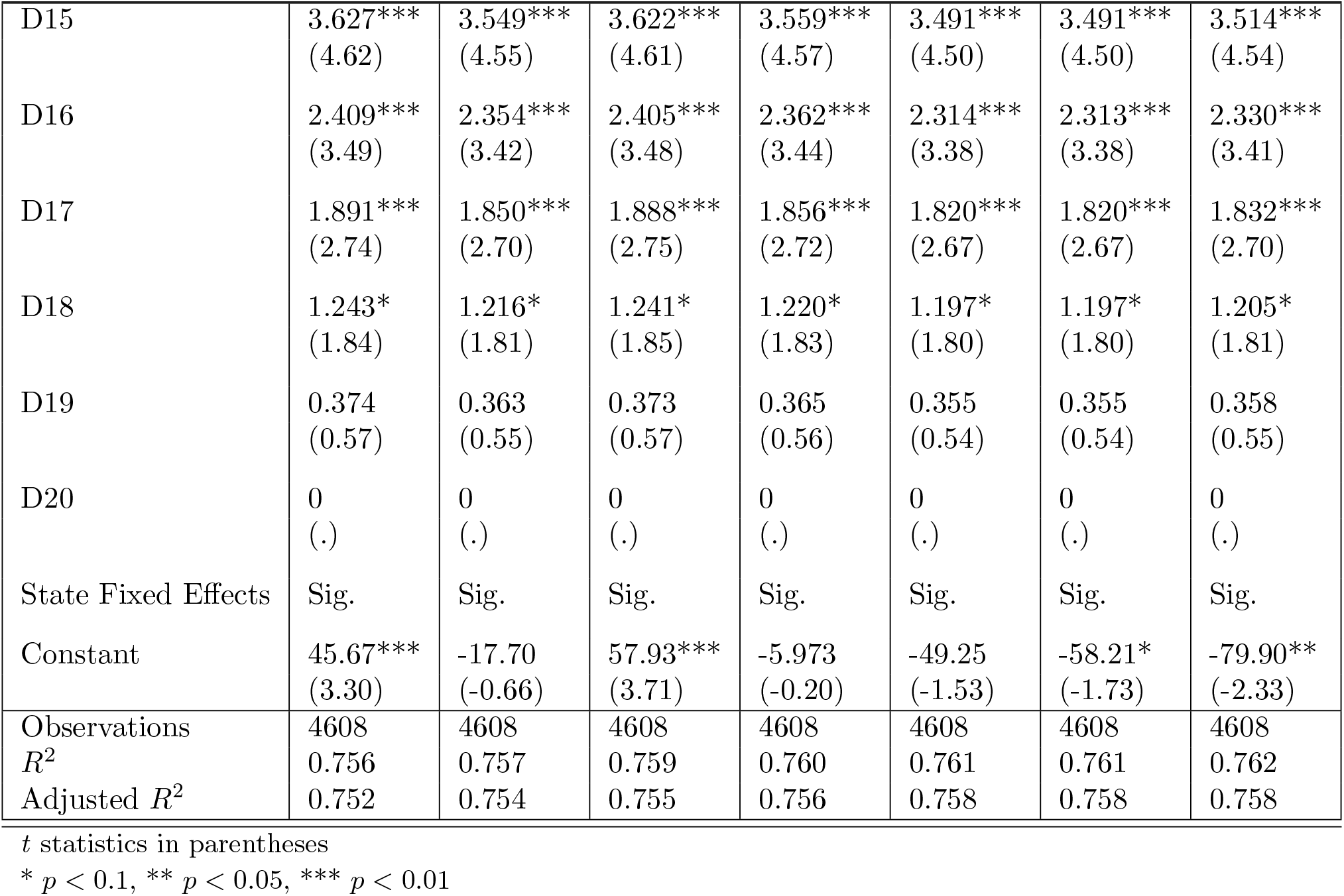
Regression to Explain Distribution of Deaths

| Variable | Model 1 | Model 2 | Model 3 | Model 4 | Model 5 | Model 6 | Model 7 |
| --- | --- | --- | --- | --- | --- | --- | --- |
| Cases Prior 14 Days<br>Per 100000 People | 0.121***<br>(12.78) | 0.119***<br>(12.69) | 0.120***<br>(12.64) | 0.119***<br>(12.70) | 0.118***<br>(12.59) | 0.118***<br>(12.59) | 0.118***<br>(12.67) |
| **Economic Variables** |  |  |  |  |  |  |  |
| ln Median Income | -7.614***<br>(-5.14) | -4.524***<br>(-2.63) | -9.285***<br>(-6.11) | -6.175***<br>(-3.75) | -1.365<br>(-0.64) | -0.685<br>(-0.31) | 0.613<br>(0.28) |
| Unempl Rate | -0.169<br>(-1.00) | -0.101<br>(-0.63) | 0.0376<br>(0.25) | 0.0846<br>(0.56) | -0.399**<br>(-2.51) |  |  |
| Gini |  | 46.38**<br>(2.44) |  | 43.61**<br>(2.21) | 58.29***<br>(2.85) | 58.62***<br>(2.85) | 66.15***<br>(3.12) |
| **Demographic Variables** |  |  |  |  |  |  |  |
| ln Population Density | 1.635***<br>(3.18) | 1.516***<br>(2.94) | 1.505***<br>(2.77) | 1.401***<br>(2.58) | 1.431***<br>(2.98) | 1.487***<br>(3.11) | 1.549***<br>(3.27) |
| % Population Over 65 | 0.446***<br>(5.53) | 0.417***<br>(5.79) | 0.394***<br>(5.43) | 0.383***<br>(5.58) | 0.370***<br>(5.87) | 0.358***<br>(5.73) | 0.462***<br>(6.46) |
| % White | -0.0794**<br>(-2.38) | -0.0654**<br>(-2.12) |  |  | -0.0110<br>(-0.37) | -0.00178<br>(-0.06) | 0.0109<br>(0.33) |
| % Asian |  |  | 0.653***<br>(5.84) | 0.704***<br>(5.76) |  |  |  |
| % Black |  |  | 0.0796**<br>(2.00) | 0.0590<br>(1.58) |  |  |  |
| % Hispanic |  |  | 0.0167<br>(0.66) | 0.0584*<br>(1.75) |  |  |  |
| % Native American |  |  | 0.0550**<br>(1.97) | 0.0455<br>(1.63) |  |  |  |
| % Multiple Race |  |  | -0.813***<br>(-3.72) | -0.692***<br>(-3.17) |  |  |  |
| % Foreign Born | -0.250***<br>(-6.04) | -0.295***<br>(-6.13) | -0.317***<br>(-3.52) | -0.440***<br>(-4.05) | -0.244***<br>(-4.90) | -0.242***<br>(-4.86) | -0.209***<br>(-4.16) |
| **Community Health/Lifestyle Variables** |  |  |  |  |  |  |  |
| Air pollution |  |  |  |  | 0.849***<br>(3.83) | 0.795***<br>(3.60) | 0.727***<br>(3.30) |
| % Flu vaccination |  |  |  |  | -0.293***<br>(-4.41) | -0.292***<br>(-4.40) | -0.302***<br>(-4.44) |
| % Diabetes |  |  |  |  | 0.481**<br>(2.49) | 0.468**<br>(2.45) |  |
| % Adult Obesity |  |  |  |  |  |  | 0.321***<br>(3.76) |
| Primary Care<br>Physicians Per Capita | -3.375***<br>(-5.84) | -4.811***<br>(-5.03) | -3.880***<br>(-6.31) | -5.111***<br>(-5.24) | -3.892***<br>(-4.65) | -3.775***<br>(-4.61) | -3.630***<br>(-4.41) |

Table 4 – continued from previous page
| Variable | Model 1 | Model 2 | Model 3 | Model 4 | Model 5 | Model 6 | Model 7 |
| --- | --- | --- | --- | --- | --- | --- | --- |
| D15 | 3.627***<br>(4.62) | 3.549***<br>(4.55) | 3.622***<br>(4.61) | 3.559***<br>(4.57) | 3.491***<br>(4.50) | 3.491***<br>(4.50) | 3.514***<br>(4.54) |
| D16 | 2.409***<br>(3.49) | 2.354***<br>(3.42) | 2.405***<br>(3.48) | 2.362***<br>(3.44) | 2.314***<br>(3.38) | 2.313***<br>(3.38) | 2.330***<br>(3.41) |
| D17 | 1.891***<br>(2.74) | 1.850***<br>(2.70) | 1.888***<br>(2.75) | 1.856***<br>(2.72) | 1.820***<br>(2.67) | 1.820***<br>(2.67) | 1.832***<br>(2.70) |
| D18 | 1.243*<br>(1.84) | 1.216*<br>(1.81) | 1.241*<br>(1.85) | 1.220*<br>(1.83) | 1.197*<br>(1.80) | 1.197*<br>(1.80) | 1.205*<br>(1.81) |
| D19 | 0.374<br>(0.57) | 0.363<br>(0.55) | 0.373<br>(0.57) | 0.365<br>(0.56) | 0.355<br>(0.54) | 0.355<br>(0.54) | 0.358<br>(0.55) |
| D20 | 0<br>(.) | 0<br>(.) | 0<br>(.) | 0<br>(.) | 0<br>(.) | 0<br>(.) | 0<br>(.) |
| State Fixed Effects | Sig. | Sig. | Sig. | Sig. | Sig. | Sig. | Sig. |
| Constant | 45.67***<br>(3.30) | -17.70<br>(-0.66) | 57.93***<br>(3.71) | -5.973<br>(-0.20) | -49.25<br>(-1.53) | -58.21*<br>(-1.73) | -79.90**<br>(-2.33) |
| Observations | 4608 | 4608 | 4608 | 4608 | 4608 | 4608 | 4608 |
| $R^2$ | 0.756 | 0.757 | 0.759 | 0.760 | 0.761 | 0.761 | 0.762 |
| Adjusted $R^2$ | 0.752 | 0.754 | 0.755 | 0.756 | 0.758 | 0.758 | 0.758 |

$t$ statistics in parentheses
\* $p < 0.1$ , \*\* $p < 0.05$ , \*\*\* $p < 0.01$

Unlike the estimation of cases reported in Table 1, a 14-day lag is used from infection to death. The results show that the number of deaths is positively related to the number of cases reported 14 days prior. The indicator variables for the days of the analysis, *D*15 through *D*19 show that relative to the 20th day (*D*20), days 15 through 18 had significantly higher deaths indicating that the rise in deaths was slowing down over time. The pandemic peaked around April 23 before its current resurgence.

The economic, demographic, and health outcomes of people are correlated. Models 1 through 7 differ in terms of the combinations of the variables that are included. Models 1 and 2 focus on the economic variables and include only one race and one health-related variable. Models 3 and 4 focus on the racial composition of the population and report results with and without the income inequality variable. Models 5 through 7 reduce the race-related factors and focus on healthcare factors.

The results of the regression as they relate to the economic variables show that unlike the spread of the virus, deaths and median household income are negatively related. The Gini coefficient of income inequality is positive and significant. Consequently, places with lower income and higher income inequality had higher fatality rates. This is consistent with our general understanding that people at lower income levels are more vulnerable to serious health shocks. Once income is accounted for, the unemployment rate and other measures of economic condition are not found to exert any additional effect on deaths.

The demographic variables show that counties with a higher concentration of people above the age of 65 had a significantly higher number of deaths, consistent with what is known about the elevated risk of the elderly to Covid-19 fatality. Models 1 and 2 that only include an indicator for the percentage of Whites in the population show that counties with a higher concentration of Whites had fewer deaths. Once the racial distribution of the non-White population is further disaggregated, the results show that counties with higher concentrations of Asians, Blacks, and Native Americans reported higher deaths. The relationship is negative for counties with more multi-racial populations and immigrants. Once the income inequality measure is included (Model 4), the impact of Blacks and Native Americans is diminished.

In all of the models reported in Table 4, access to health care in terms of the number of primary care physicians per capita is included. This variable is significant and negative in every model showing that access to physicians to take care of people’s everyday health needs is a protection against the most severe outcome of the disease. Consistent with this finding are the results that show that places with higher concentrations of populations with high-risk health outcomes such as obesity and diabetes have higher reported deaths. Using county-level data, papers such as Wu (2020) demonstrate that air pollution is a risk factor for Covid-19 death. The results reported here support their finding. The health results also show that counties with higher rates of flu vaccination have a lower risk of death from the virus. It is to be noted that ours is an observational study, so this result by no means implies that the flu vaccine is a protection against the virus. Rather, the result is to be interpreted in terms of health awareness and action. Counties with higher percentages of the population that get vaccinated against the flu are perhaps places where people, in general, are more responsive to threats to their health and take steps to mitigate them. It may reflect their overall approach to their health and wellbeing.

The results show that the economic factors are important for explaining the differential impacts experienced by counties across the country both in terms of confirmed cases and deaths reported. The demographic and health-related factors are more pronounced in the estimation of deaths than reported cases. This is not surprising since the virus does not discriminate based on any factor other than immunity, but the severity of the disease that can lead to a fatal outcome depends on underlying health and demographic factors.

## 4 Conclusion

This paper has examined the differential experience of infections and deaths across the US due to the COVID-19 pandemic. The analysis of the number of cases is based on an epidemiological model in which we included a county fixed effect. This is a novel way to introduce heterogeneity in such a model. A dynamic panel regression of the number of cases included the potential number of interactions between susceptible and infected individuals as a proportion of the population along with county fixed effects. The results of the model were used to construct a Vulnerability Index for each county. Economic, demographic, and health/lifestyle factors were used to explain the differences in the Vulnerability Index across the counties. The results showed that counties with higher economic activity have higher vulnerability. The results show that regions around international airports experienced higher numbers of cases than ones that are over 300 miles away. This is consistent with the fact that the virus has arrived on the US shores through travelers coming to the US from abroad. Counties with more elderly and non-White populations are more vulnerable and so are counties with higher income inequality and housing problems.

The results related to deaths show that counties with lower income and higher cases experienced higher deaths. It is to be noted that counties with higher income reported more cases but when it came to fatality, lack of income is a risk factor. Counties with higher population density and higher income inequality also experienced more deaths. Counties with higher percentages of non-Hispanic Blacks, Native Americans, and Asians more likely to die relative to counties with non-Hispanic Whites. Counties with more personal care physicians per capita experienced lower deaths, and so did counties with a lower percentage of the population with health-risk factors such as obesity and diabetes. Air pollution is also found to be associated with higher deaths. While studies show that long term exposure to air pollution can cause long term vulnerability to lung-related diseases, it is to be noted in an observational study such as this one should not draw conclusions about health-risk factors from observed results. This is true about the result related to flu vaccination as well. The results suggest that counties with more health-conscious populations that take preventive actions have a better outcome in terms to surviving the Covid-19 disease.

The economics literature is not extensive in the area of pandemics and epidemics in developed countries. The contribution of this study is to understand the various socioeconomic conditions that can make a county or region more vulnerable to both disease spread and severity of cases. A national strategy to prepare the infrastructure for controlling the spread of infectious diseases should consider these factors and develop Vulnerability Indexes for regions across the country.

## Data Availability

All data used in the project are available in the files stored in the link below.

https://github.com/nivedita-mukherji/Covid-socioeconomic-research-project

## Footnotes

1 Different specifications used in estimating the index led to similar rankings of the index even if the actual values of the coefficients varied. Since it is the relative rankings that matter for the analysis of the underlying socioeconomic factors, the results of the following analysis are largely insensitive to the exact specifications used to derive the index values.

2 The diagonal values of the weight matrix used for the calculation of the weighted international passengers are zeros. Consequently, the weighted values measure the impact in the surrounding areas only.

